# Causality of Genetically Determined Monounsaturated Fatty Acids on Risk of Cardiovascular Disease: A Mendelian Randomization Study

**DOI:** 10.1101/2023.09.06.23295142

**Authors:** Danial Habibi, Mahdi Akbarzadeh, Farshad Teymoori, Sahand Tehrani Fateh, Sajedeh Masjoudi, Amir Hossein Saeidian, Farhad Hosseinpanah, Noushin Mosavi, Hakon Hakonarson, Fereidoun Azizi, Soleymani T Alireza, Mehdi Hedayati, Maryam Sadat Daneshpour, Marjan Mansourian

**Affiliations:** Department of Biostatistics and Epidemiology, School of Health, and Student Research Committee, School of Health, Isfahan University of Medical Sciences, Isfahan, Iran; Cellular and Molecular Endocrine Research Center, Research Institute for Endocrine Sciences, Shahid Beheshti University of Medical Sciences, Tehran, Iran; Nutrition and Endocrine Research Center, Research Institute for Endocrine Sciences, Shahid Beheshti, Tehran, Iran; School of Medicine, Tehran University of Medical Sciences, Tehran, Iran; Center for Applied Genomics, The Children’s Hospital of Philadelphia, Abramson Research Building, Suite 1016I, 3615 Civic Center Boulevard, Philadelphia, PA, 19104-4318, USA; Obesity Research Center, Research Institute for Endocrine Sciences, Shahid Beheshti University of Medical Sciences, Tehran, Iran; Department of Metbolism, digestion and reproduction, Imperial College London; Department of Pediatrics, University of Pennsylvania Perelman School of Medicine, Philadelphia, PA, USA; Division of Human Genetics, The Children’s Hospital of Philadelphia, Philadelphia, PA, USA; Endocrine Research Center, Research Institute for Endocrine Sciences, Shahid Beheshti University of Medical Sciences, Tehran, Iran; International Artificial Intelligence Department, School of Management, National Yunlin University of Science and Technology, Douliu, Taiwan; Epidemiology and Biostatistics Department, School of Health, Isfahan University of Medical Sciences, Isfahan, Iran

**Keywords:** Serum monounsaturated fatty acids, cardiovascular diseases, angina, atherosclerotic, ischemic heart disease, myocardial infarction, high blood pressure, Mendelian randomization, Genome-wide Association study

## Abstract

**Background/Aim:** The possible association between serum monounsaturated fatty acids (MUFAs) and the risk of cardiovascular diseases (CVDs) has been examined in observational studies, which indicate controversial findings. In the current study, we used the Mendelian randomization (MR) analysis to determine the causal relationship of genetically determined serum MUFAs with the risk of various CVD outcomes, including angina, atherosclerotic, ischemic heart disease (IHD), myocardial infarction (MI), and high blood pressure (BP).

**Method:** The summary statistics dataset on the genetic variants related to serum MUFAs was used from the published GWAS of European descent in UK Biobank participants (N=114,999). Genetic variants underlying angina, atherosclerotic, IHD, MI, and BP events were ascertained using a GWAS dataset of 461,880 (case= 14,828, control= 447,052), 463,010 (case= 12,171, control= 450,839), 361,194 (case= 20,857, control= 340,337), 462,933 (case= 10,616, control= 452,317), and 461,880 (case= 124,227, control= 337,653) European descent participants from the UK Biobank, respectively.

**Results:** Our results showed that MUFAs were associated with angina [OR_IVW_= 1.005, 95% CI: 1.003– 1.007, p = <0.001; Cochran’s Q=23.89, p=0.717, I^2^=0.0%; Egger intercept= -0.0003, p=0.289], atherosclerotic [OR_IVW_= 1.005, 95% CI: 1.003–1.007, p = <0.001; Cochran’s Q=42.71, p=0.078, I^2^=27.4%; Egger intercept= -0.0004, p=0.146], IHD [OR_IVW_= 1.004, 95% CI: 1.001–1.007, p = 0.005; Cochran’s Q=42.75, p=0.172, I^2^=18.1%; Egger intercept= -0.0001, p=0.827], MI [OR_IVW_= 1.001, 95% CI: 0.999– 1.003, p = 0.199; Cochran’s Q= 23.03, p=0.631, I^2^=0.0%; Egger intercept= -0.0003, p=0.196], and BP [OR_WM_= 1.008, 95% CI: 1.001–1.015, p = 0.022; Cochran’s Q= 52.87, p=0.015, I^2^= 37.6%; Egger intercept= 0.0002, p=0.779]. These results remained consistent using different Mendelian randomization methods and sensitivity analyses.

**Conclusion:** In the present MR analysis, serum MUFA levels were associated with the risk of angina, atherosclerotic, IHD, MI, and BP. These findings prompt significant questions about the function of MUFAs in the progression of CVD events. Further research is required to elucidate the connections between MUFAs and CVD to contribute to health policy decisions in reducing CVD risk.

## Introduction

Fatty acids are a constituent of a group of carboxylic acids comprising saturated and unsaturated fatty acids (1). Saturated fatty acids lack double bonds, while monounsaturated fatty acids (MUFA) and polyunsaturated fatty acids (PUFAs) have one double bond or more, respectively (2).

The National Institute of Medicine (NIH), the United States Department of Agriculture (USDA), the European Food and Safety Authority (EFSA), and the American Diabetes Association (ADA) have not provided appropriate guidelines regarding MUFAs consumption (3). In contrast, the Academy of Nutrition and Dietetics recommends 15% to 20% MUFA of daily total energy expenditure (4), and the American Heart Association limited the consumption of MUFAs by 20% MUFA in their dietary guidelines (3). Additionally, nutritionists and physicians highly recommend the Mediterranean Diet for its health-promoting effects, which is rich in MUFA (5).

MUFA may influence various markers associated with CHD, including postprandial vascular function, vascular function markers, and serum lipid and lipoprotein profile (6–9). MUFA-enriched foods (e.g., plant-based oils) have also been recommended to reduce the risk of cardiovascular diseases, body weight, and other inflammation-related diseases (10,11).

Previous studies examining the association between MUFA consumption and CVD risk have faced limitations and yielded inconclusive results (12–17). The varying associations between MUFA intake from plant and animal sources with CHD risk indicate that plant-based foods are preferable sources of MUFAs for preventing CHD (18). Moreover, the original research findings may not be entirely comparable since the analysis did not simultaneously consider the macronutrients with different health effects (19). However, the findings on the relationship between MUFA and CVD-related traits are based on observational studies, and the lack of a clear intuition is comprehensibly evident. So, the knowledge gap is whether MUFA can be considered a causal factor for CVD-related traits. Therefore, we aim to investigate this hypothesis through a Mendelian randomization (MR) study.

## Methods Study Design

A two-sample MR study design was used, in which summary data from genome-wide association studies (GWAS) provide a rich resource of exposure–disease pathways (20). To estimate causal inference, we selected MUFA as exposure and angina, atherosclerotic heart disease, ischemic heart disease, heart attack, and high blood pressure as outcomes from European ancestry (Table 1 and Supplementary Figure S1).

**Table 1.** Detailed information of data used for analyses.

| NAME | GWAD ID | Sample size | Role |
| --- | --- | --- | --- |
| Angina | ukb-b-8468 | 461,880 (case= 14,828, control= 447,052) | Outcome |
| Atherosclerotic | ukb-b-1668 | 463,010 (case= 12,171, control= 450,839) | Outcome |
| Ischemic heart disease | ukb-d-I9-IHD | 361,194 (case= 20,857, control= 340,337) | Outcome |
| Myocardial infarction | ukb-b-15829 | 462,933 (case= 10,616, control= 452,317) | Outcome |
| High blood pressure | ukb-b-14177 | 461,880 (case= 124,227, control= 337,653) | Outcome |
| Monounsaturated Fatty Acids | met-d-MUFA | 114,999 | Exposure |

### Genetic Variants of Exposure

We used summary GWAS information of the European ancestry on genetic variants associated with MUFA and cardiovascular diseases in figure 1. To specify the genetic instruments, we should require a genome-wide threshold of P < 5 × 10^−8^, while this threshold could be relaxed in instances with less than 15 independent loci (21). Among all the identified variants, only single nucleotide polymorphisms (SNPs) were significantly associated with MUFA in P < 5 × 10^−6^ (22). To minimize correlations between the SNPs, we conducted an LD-clumping threshold of r^2^ < 0.2, in which SNPs with the lowest p-value are retained(22,23).

**Figure1:**
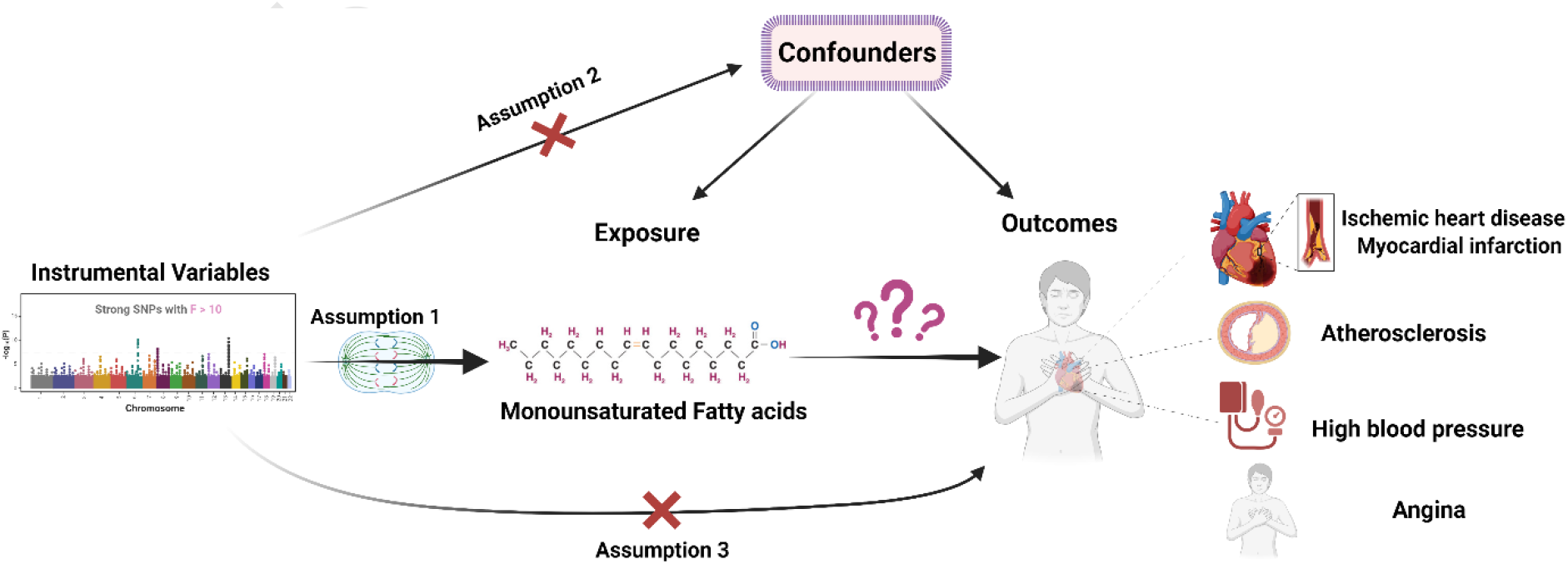
causal association between monounsaturated fatty acids and risk of cardiovascular disease.

For SNPs to be valid, they should be associated with the exposure, irrelevant to confounders of the exposure-outcome association; they should also affect the outcome only through the exposure (24). The F-statistic was calculated to test the weak IV bias to avoid weak instrumental bias. If the F-statistics>10, the strength of the SNP selected was strong (25). We applied the PhenoScanner (http://www.phenoscanner.medschl.cam.ac.uk/) to assess whether the selected SNPs were associated with other traits at genome-wide significance levels (26,27). Then, after removing the identified pleiotropic SNPs, we rerun the results. The relevant information was extracted, including SNP, effect allele, non-effect allele, effect allele frequency, effect sizes, standard error, sample size, and p-value.

The palindromic SNPs with intermediate allele frequencies from 0.01 to 0.30 were disregarded from the above-selected SNPs. Additionally, SNPs with a minor allele frequency (MAF) less than 0.01 were omitted to avoid potential statistical bias from the original GWAS since they usually carry low confidence (28).

## Statistical analysis

The inverse variance-weighted (IVW) with a multiplicative random-effects model was performed to primarily analyze the causal associations between exposure to monounsaturated fatty acids and outcome (cardiovascular diseases) (29). This method might be affected by pleiotropy or invalid instrument bias; we assessed the validity and robustness of the results by executing several sensitivity analyses, including MR-Egger, weighted median, simple mode, and weighted mode method (30). Moreover, we applied several other approaches, including the Maximum Likelihood Estimation (MLE) method, Robust Adjusted Profile Score (RAPS), MR-Lasso, constrained maximum likelihood, mode-based and debiased inverse variance weighted to detect the robustness of our results (31). MR Steiger test was performed to assess the direction of causation and whether the observed associations were directionally causal (P < 0.05 was defined as statistically significant) (32).

Cochran’s Q statistic and the I^2^ index for MR-inverse-variance weighted analyses were used to detect heterogeneity and pleiotropy, and Rucker’s Q statistic for MR-Egger (33). We used the MR-Egger method (by intercept tests) to evaluate horizontal pleiotropy (34). Cook’s distance and Studentized residuals to ascertain whether any individual SNPs were detected as outliers and influential points in driving the analysis results (34). Moreover, we used MR-PRESSO and RadialMR (using Cochran’s Q-statistic) to identify outliers with potential pleiotropy.

Leave-one-SNP-out analysis and its plot were conducted to assess the influence of potentially pleiotropic SNPs on the causal estimates by systematically removing one SNP at a time (35). Funnel plot depicted to detect directional pleiotropy whether causal estimates from weaker variants tend to be skewed in one direction (36). We drew a forest plot of Mendelian randomization for the association between genetically predicted monounsaturated fatty acids and cardiovascular disease. We also demonstrated a scatter plot to inspect outliers, genetic association, and causal estimates visually. The process of MR analyses and the results are publicly available through the following HTML link: https://akbarzadehms.github.io/MUFACVD-MR/

The principal statistical analyses was calculated with the help of R software (version 4.0.3) by “TwoSampleMR,” “MendelianRandomization”, “MR-PRESSO,” “RadialMR”, and “mr.raps” packages (37–39), and mrrobust Stata package (40). Results were reported as odds ratios (OR) with corresponding 95% CI. To account for multiple testing in monounsaturated fatty acids concerning the five outcomes, we used a Bonferroni-corrected threshold of P < 0.01 (α = 0.05/5 outcomes). Associations with P-values between 0.01 and 0.05 were regarded as suggestive evidence of associations.

## Results

### SNPs selection

From 10,331,678 SNPs of monounsaturated fatty acids, we reached 44 SNPs after P<5×10^−6^ and LD clumping (Supplementary sheet1). All F-statistics of SNPs were greater than 10, which elucidated no weak instrument bias (Supplementary sheet1). In the harmonizing step (action=2), after removing SNPs duo to palindromic, the PhenoScanner tool, and potential outliers and influential points, we attained 28, 32, 36, 27, and 34 SNPs for angina, atherosclerotic, ischemic heart disease, myocardial infarction, and hypertension, respectively (figure2).

**Figure 2:**
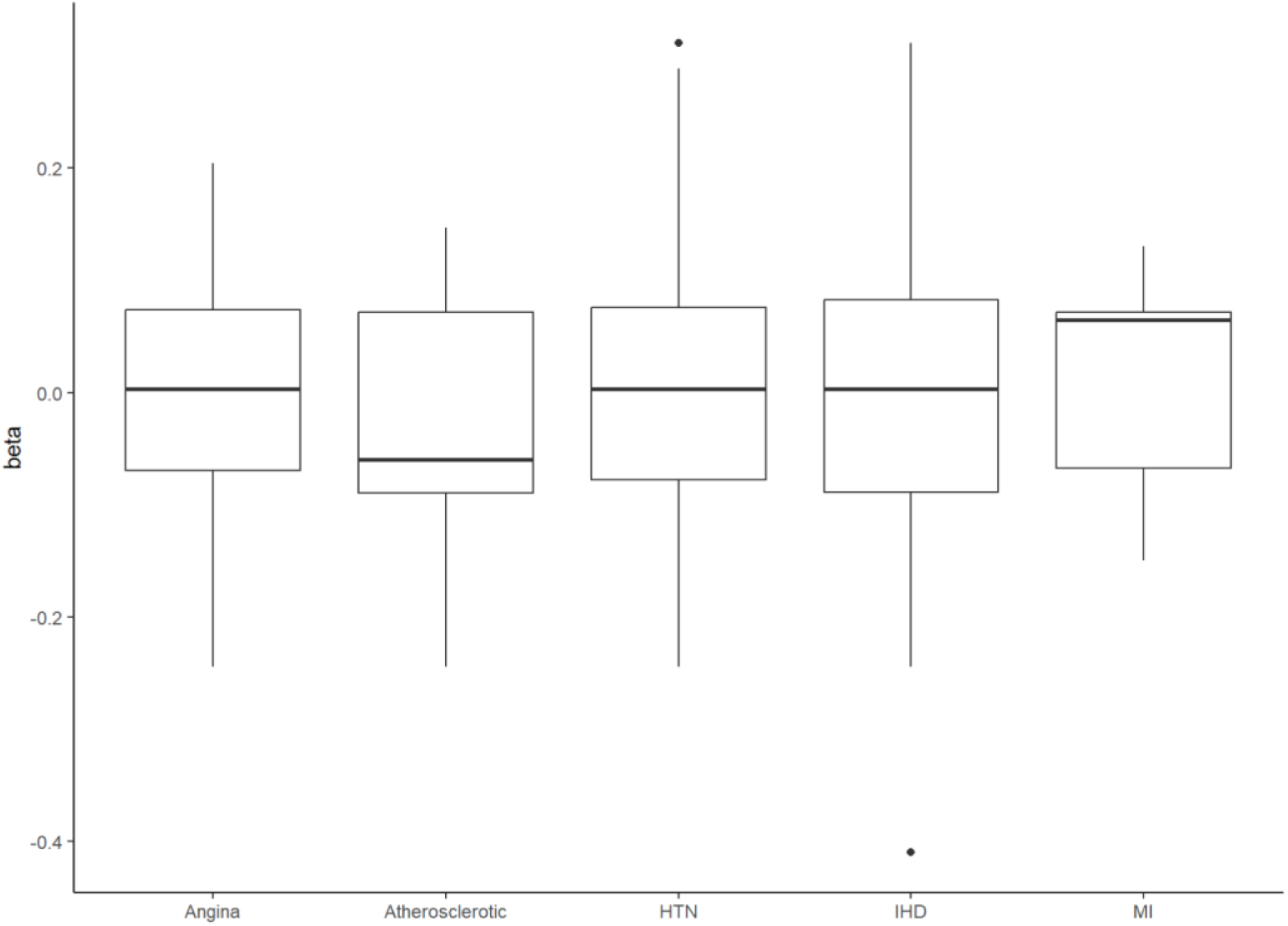
The visualized results of the selected SNPs using bar charts of the mean and standard deviation into each CVD. HTN: Hypertension, IHD: Ischemic heart disease, MI: Myocardial infarction.

To roughly display relationship of the undefined causal variables to CVD disease of study, we used beta of MUFA regressed on the undefined causal variable versus the CVD disease. All coefficients indicated positive association between MUFA and CVD (Supplementary Figure S2).

### Mendelian Randomization analysis

The inverse variance weighted method showed that a positive association could exist between monounsaturated fatty acids and angina, atherosclerotic, ischemic heart disease, myocardial infarction, and hypertension (figure3).

**Figure 3:**
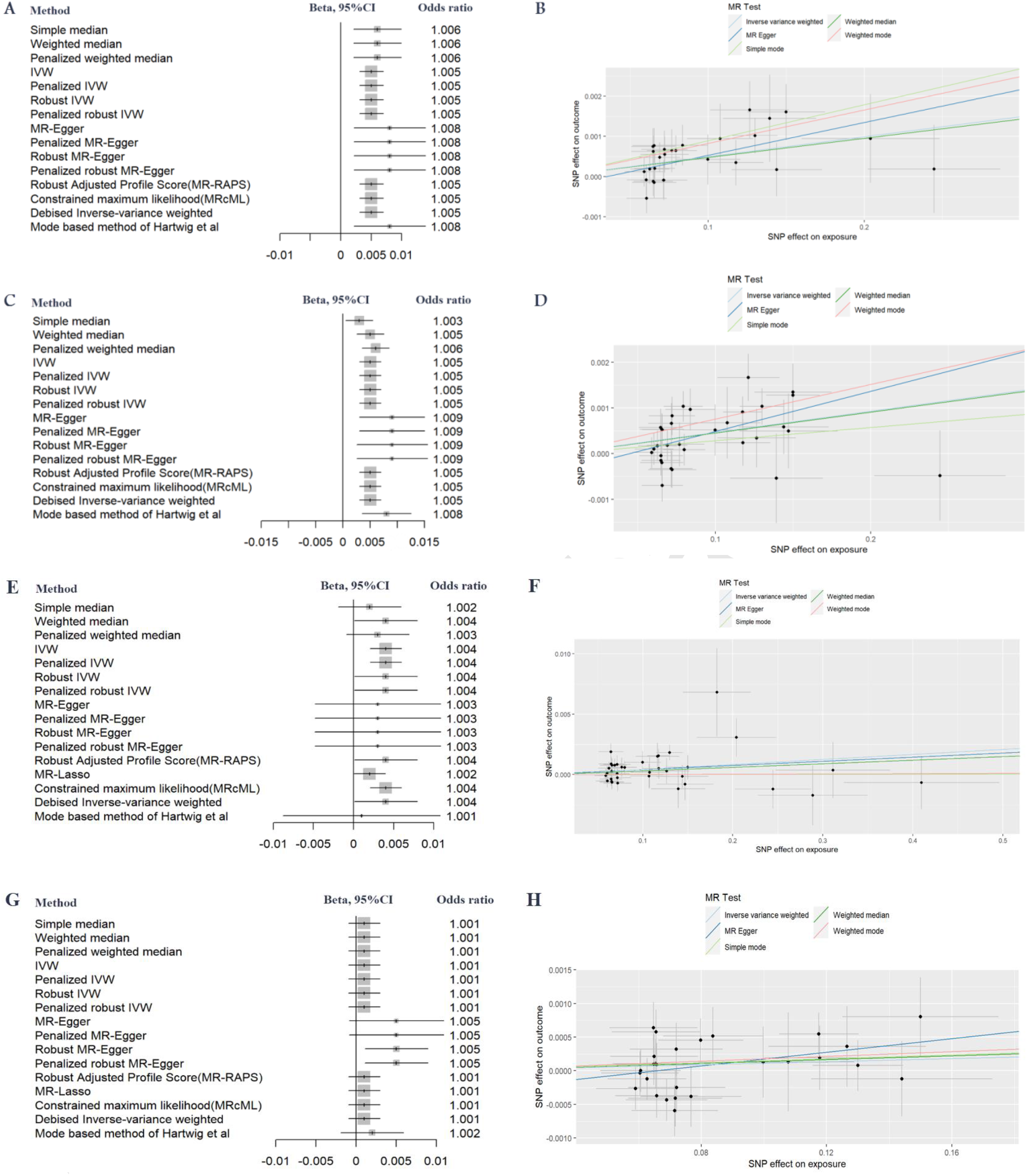
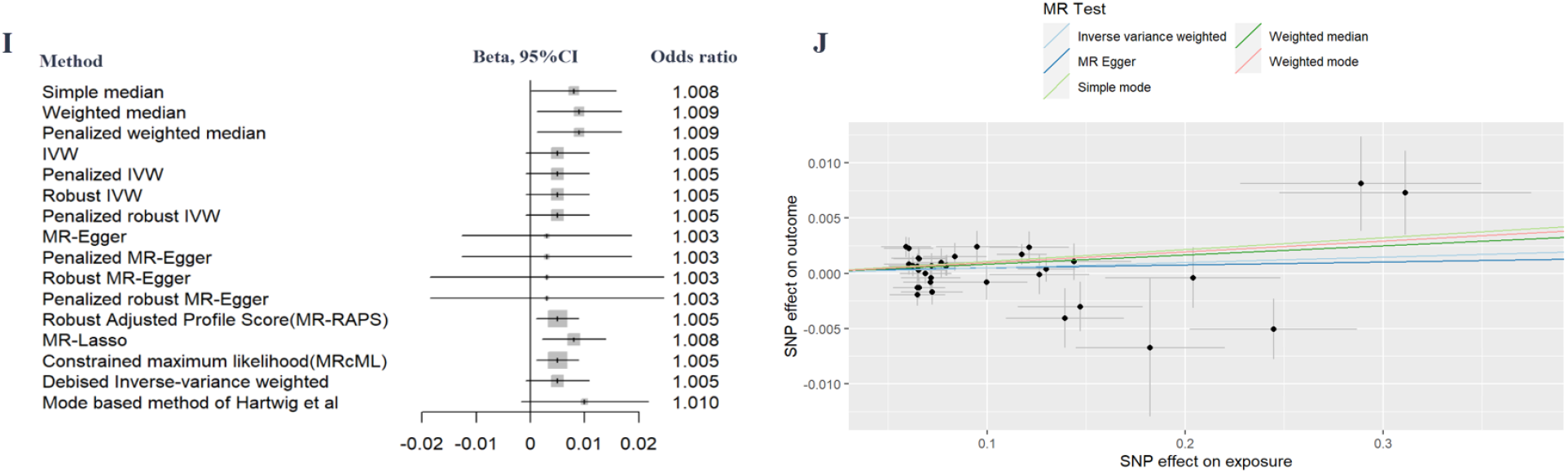
MR analysis. A and B: angina; C and D: atherosclerotic; E and F: IHD; G and H: Myocardial infarction; I and J: Hypertension. Left plots (A, C, E, G, I) shows forest plot of difference methods, and right plots (B, D, F, H, J) exhibits scatter plot based on main methods (inverse variance weighted, weighted median, MR Egger, weighted mode, and simple mode).

After assessing the heterogeneity pleiotropy test as well as potential outliers and influentials SNPs according to Cook’s distance (rs10152355, rs591592, rs77548999, and rs1860356), angina result showed OR_IVW_= 1.005, 95% CI: 1.003–1.007, p = <0.001; Cochran’s Q=23.89, p=0.717, I^2^=0.0%; Egger intercept= -0.0003, p=0.289 (Supplementary sheet2).

For atherosclerotic after evaluating the heterogeneity pleiotropy test as well as potential outliers and influentials SNPs according to RadialMR (rs2642636, rs429358, and rs56289821), represented OR_IVW_= 1.005, 95% CI: 1.003–1.007, p = <0.001; Cochran’s Q=42.71, p=0.078, I^2^=27.4%; Egger intercept= -0.0004, p=0.146 (Supplementary sheet3).

For IHD, after evaluating the heterogeneity pleiotropy test as well as potential outliers and influentials SNPs according to Cook’s distance (rs12185072, rs174418, and rs115849089), represented OR_IVW_= 1.004, 95% CI: 1.001–1.007, p = 0.005; Cochran’s Q=42.75, p=0.172, I^2^=18.1%; Egger intercept= -0.0001, p=0.827 (Supplementary sheet4).

For Myocardial infarction, after evaluating the heterogeneity pleiotropy test as well as potential outliers and influential SNPs according to Cook’s distance (rs61999891, rs174418, and rs115849089), indicated OR_IVW_= 1.001, 95% CI: 0.999–1.003, p = 0.199; Cochran’s Q= 23.03, p=0.631, I^2^=0.0%; Egger intercept= -0.0003, p=0.196 (Supplementary sheet5).

For BP, after evaluating the heterogeneity pleiotropy test as well as potential outliers and influential SNPs according to Cook’s distance (rs12185072, rs174418, and rs115849089), showed OR_WM_= 1.008, 95% CI: 1.001–1.015, p = 0.022; Cochran’s Q= 52.87, p=0.015, I^2^= 37.6%; Egger intercept= 0.0002, p=0.779 (Supplementary sheet6).

### Sensitivity analyses

According to the results of Cochran’s Q test, only statistically significant heterogeneity was observed in BP. MR-Egger regression intercept analysis revealed no directional horizontal pleiotropy in all cardiovascular diseases (Supplementary Figure S8).

MR pleiotropy residual sum and outlier test (MR-PRESSO) methods, MR pleiotropy residual sum and outlier (Radial MR), and Cook’s distance were used to assess the potential of the presence of and the outlying SNPs and reassess the effect estimates (Supplementary Figure S3). Scatter plots (Supplementary figure S4) and leave-one-out plots (Supplementary Figure S5) were visually examined to test the influence of outlying values.

Moreover, we depicted funnel and forest plots of causal association between monounsaturated fatty acids on cardiovascular diseases (Supplementary Figures S7 and S8). The weak instrumental variable was evaluated through the F-statistic (Supplementary sheet1). We also checked whether SNPs within the HLA region existed in our MR analysis (Supplementary sheet1).

## Discussion

As the first one, the current study has examined the dose-response causal association between serum MUFAs and the risk of various CVD outcomes, including angina, atherosclerotic, IHD, MI, and high BP, using MR analyses with large GWAS summary statistics. The present MR analysis showed that genetically higher serum levels of MUFAs can be related to increased risk of CVDs, including angina, atherosclerotic, IHD, MI, and elevated BP.

The positive causal relationship reported in the present study is in line with the findings of several previous observational studies that supported the hypothesis of the unfavorable impact of high serum MUFAs levels in the occurrence of CVD events(41–43). Evidence supported by our MR analysis is in agreement with the results of a meta-analysis of five UK-population-based cohorts, and one matched case-control study that suggested that high levels of circulating MUFAs, such as palmitoleic acid and oleic acid, were related to a higher risk of coronary heart disease and stroke(41). Also, Akbaraly *et al*. showed the concordance between higher levels of serum MUFAs associated with low adherence to a healthy diet determined by the healthy eating index. They reported a positive association between high levels of MUFAs and an increased risk of CVD in the population of n the Whitehall II study(42). Furthermore, an observational study conducted on CKD patients revealed that elevated serum MUFA levels can be associated with an increased risk of CVDs, and which has been claimed that these high levels of MUFAs are due to high endogenous MUFA synthesis(43). However, contrary to the results of our study, a systematic review and meta-analysis reported that there was no significant association between dietary or circulating MUFAs and the risk of coronary diseases(17).

Prior research has also explored the relationship between the serum level of some MUFAs individually and CVD risk, which has shown conflicting findings. A prospective study in the framework of the Multi-Ethnic Study of Atherosclerosis revealed that high circulating oleic acid levels may be a predictive risk factor for the incidence of CVD events and mortality(44); also, a US cohort reported that there was a significant positive relationship between plasma monounsaturated fatty acids, especially palmitoleic acid and risk of ischemic stroke(45). Furthermore, in two other studies, the high serum level of palmitoleic acid was presented as an index for increasing the risk of coronary heart disease(46) and coronary artery disease(47). However, other studies suggested that oleic acid was related to a lower risk of acute MI(48) and stroke incidence(49). Also, a cross-sectional study conducted on US adults in the National Health and Nutrition Examination Survey showed that high circulating palmitelaidic acid was inversely linked to CVD events risk(50).

Our findings are inconsistent with the results of Mazidi *et al*. study that, based on an MR analysis, has reported genetically higher serum of some MUFAs, including 10-heptadecenoate, myristoleic, oleic, and palmitoleic acids, may not be associated with the risk of some CVD events, such as coronary heart disease, MI, cardioembolic stroke, and ischemic stroke(51). The reasons for the controversy in our findings with the results of the previous study can be that, first, in the Mazidi *et al*., only some MUFAs were examined in relation to the risk of CVDs, while in the present study, we determined the association of total serum MUFAs with CVD events risk; Considering that for the occurrence of the possible connection between serum MUFAs levels and CVD risk, the most important effective factor is the high total vascular concentration of these fatty acids, which can be a predictor of the accumulation and adhesion of MUFAs in the vessels, the creation of vascular stiffness, and susceptibility to the occurrence of CVD events. Finding out the possible relationship between serum MUFAs and the mentioned clinical abnormalities is more likely in our study, which focuses on the total serum MUFA concentration. It is also necessary to mention that the types of CVD outcomes examined in our study have differences from the CVD events assessed in the Mazidi *et al*. study, Considering that the metabolic and biological abnormalities that are responsible for the occurrence of CVDs may be different based on the type of CVD outcomes that examined in two these studies. Also, the direction and intensity of the serum MUFAs effect on these various CVD events can differ. Therefore, based on these explanations, the inconsistency observed in the results of these two studies can be justified.

Serum MUFAs, as a part of the lipid profile, are highly dynamic molecules that can influence a broad spectrum of cell signaling pathways, impacting lipid metabolism(41,52), blood pressure(53,54), glucose homeostasis(55,56), inflammation and oxidative stress process(57), and endothelial function(58). Also, MUFAs in the fatty acids pool that circulate in the bloodstream reflect the dynamic combination of several metabolic pathways regulated by hormonal signaling. These metabolic pathways include adipose tissue lipolysis, the hydrolysis of blood triglycerides (TGs) in TG-rich lipoproteins, re-esterification to TGs within adipocytes, and peripheral utilization of fatty acids(41). Fatty acids are also subject to desaturation and elongation processes that transform them into other fatty acids(41). It should be noted isolating the metabolic effects and coronary outcomes of circulating MUFAs from other fats can be challenging; it is suggested that any unusual changes in those metabolic and biological pathways of MUFAs may have unfavorable metabolic and vascular consequences in the long term. These changes can occur when the circulation levels of free fatty acids, such as MUFAs, are excessive and undesirable. It is crucial to note that dietary intake of MUFAs does not have a significant role in determining serum levels of MUFAs. Instead, their endogenous synthesis, managed by the hepatic de novo lipogenesis process and by stearoyl CoA desaturase 1, is the primary factor in determining serum MUFA levels(41,43). Therefore, these associations may indicate an underlying activation of hepatic de novo lipogenesis, which is linked to higher levels of blood TGs, abnormal fat deposition, and insulin resistance(59). Also, the levels of circulating fatty acids can impact several vital functions of the endothelium, such as managing vascular tone, controlling blood flow and fluidity, regulating angiogenesis, and modulating inflammatory responses(58,60,61). The abnormal circulating levels of free fatty acids in the bloodstream can create an environment conducive to vascular dysfunction, encompassing a range of clinical disorders. It can cause a decrease in nitric oxide production, the production of cytokines, damage to vasodilation, an overactive platelet response, escalated inflammatory response, and oxidative stress. The resulting clinical irregularities can subsequently lead to increased CVD events(58).

Although, based on the valuable findings of the current study, high levels of serum MUFAs were linked to a greater risk of developing CVD outcomes, it would be impractical and narrow-minded to propose that dietary changes based on reducing the consumption of fats-containing food choices could effectively mitigate the risk of CVD outcomes(3). Firstly, we did not determine the separate role of important MUFAs in the blood (such as palmitoleic acid and oleic acid) in the risk of cardiometabolic disorders. In contrast, the biological role and metabolic pathways of each of these MUFAs may differ, and this issue may be practical on serum levels of various types of MUFAs and their impact on CVD outcomes incident. Also, the sensitivity and tendency of different MUFAs to different metabolic processes, such as esterification, peroxidation, and calcification, which can affect their deposition and accumulation in the vessel wall, can be different. Therefore, recommending to consume or not to consume one or more food groups that usually contain several types of unsaturated fatty acids, such as various MUFAs, may not be an effective and accurate nutritional intervention in this case. Some MUFAs, such as palmitoleic acid and oleic acid, that are in high levels in the blood circulation and are likely to be observed at a high level in biochemical measurement do not usually reflect dietary levels of these fatty acids(41,43) because these fatty acids are also synthesized endogenously, by the process known as de novo lipogenesis using non-lipid dietary precursors, including carbohydrate and alcohol intake. So, limiting the consumption of food sources containing high levels of them may not effectively reduce the serum level of these MUFAs(41,43). These points mentioned above can justify our results’ inconsistency with the previous meta-analysis’s findings, which revealed that unsaturated fatty acids cannot be a risk factor for increasing cardiovascular disorders(17).

The current study has important strengths; this study is the first study that has used a TSMR method to assess the possible causal association between serum MUFA and the risk of various CVD outcomes and provided a detailed and holistic view of the real dose-response effect of MUFAs serum levels in predicting the risk of various CVD outcomes. Another important strength of the current study was using a large sample size from the UK Biobank database, which presents a rich genetic and phenotypic data source. However, some limitations of the present study deserve to be mentioned. The GWAS summary statistics for different types of MUFAs (e.g., palmitoleic acid, oleic acid, 10-heptadecenoate, and myristoleic) were unavailable for this study. Therefore, we could not determine the causal relationship of each MUFAs separately with the risk of CVD outcomes. This limitation can be significant because different types of MUFAs may behave differently biologically in influencing the occurrence of chronic diseases. Also, the degree of their susceptibility to peroxidation, esterification, accumulation, and deposition in blood vessels differs based on the type of MUFAs, which can be effective in their predictive role for CVDs. Although the MR approach provides a causal relationship between serum MUFA levels and the risk of various CVD outcomes, it does not provide specific evidence on the mechanistic pathways justifying the relationship between exposure and outcome. Experimental studies and clinical trials that focus on the dietary MUFA intakes can help determine how and through which mechanisms the consumption of MUFAs can affect the serum levels of MUFAs and, subsequently, the risk of CVDs.

## Conclusions

Evidence from our MR analysis supports a causal association between high serum levels of MUFAs and the risk of various CVD outcomes. However, conclusions should be made with more caution due to the limitations of MR investigations and the complexity of the linkage between serum MUFAs and CVD risk. Conducting more studies in populations with different characteristics using more accurate approaches and measurement tools can help confirm the evidence extracted from this study. Also, performing clinical studies can provide underlying biological mechanisms to justify the positive causal relationship discovered between serum MUFA levels and CVD events risk.

## Declarations

## Acknowledgments

We want to acknowledge the participants and investigators who made summary data available.

## Consent for publication

Not applicable.

## Data Availability Statement

The original data used are publicly available at https://gwas.mrcieu.ac.uk/. The original contributions presented in the study are included in the article/Supplementary Material; The process of MR analyses and the results are publicly available through the following HTML link: https://akbarzadehms.github.io/MUFACVD-MR/

## Ethical approval and consent to participate

The Isfahan University of Medical Sciences ethics committee approved this study (Research Approval Code:340111 & Research Ethical Code: IR.MUI.RESEARCH.REC.1401.063).

Informed consent was obtained from all subjects in the original genome-wide association studies. The research has been performed by the Declaration of Helsinki.

## Author Contributions

DH and MA drafted the initial manuscript. MA, DH, FT conceived the study, analyzed the data, and drafted the initial manuscript. MA and DH desige the html file. DH, MA, SM, and AST and STF generated Tables and Figures. DH, MA, FH, FZ, MH, AHS, NM, HH, MSD, and MM revised the manuscript. MSD and MM supervised, edited, and finalized the manuscript. All authors reviewed and approved the final manuscript.

